# The Association Between Medicare Eligibility on Elective Procedures and Hospital Transfers: A regression discontinuity design applied to the National Inpatient Sample

**DOI:** 10.1101/2024.01.12.24301234

**Authors:** Akshay Swaminathan, Wasan Kumar, Benjamin Jacobson, Lathan Liou, Ivan Lopez, Caroline Yao, Sarthak Shah

**Author notes:** **Corresponding Author:** Akshay Swaminathan. **Funding Statement:** No funding was obtained for this study. **COI Statement:** The authors declare no competing interests. **Author Contributions:** Conceptualization: AS, WK, BJ, CC, SS Writing: AS, WK, LL, BJ Data acquisition: LL Data analysis: AS, IL, BJ Critical review: All authors. **Data availability statement:** Data used for this analysis is available from HCUP at www.hcupus.ahrq.gov/nisoverview.jsp.

## Abstract

**Introduction:** Access to affordable health coverage for individuals aged 51 and older, particularly those transitioning out of employer-sponsored insurance or with existing health conditions, is challenging in the United States. This study investigates the potential impact of lowering the Medicare eligibility age on healthcare utilization, focusing on elective procedures among late middle-aged individuals. Previous studies indicate significant increases in healthcare utilization around the Medicare eligibility threshold, but a national-level analysis is needed to understand the implications of policy changes in Medicare eligibility.

**Methods:** This retrospective cohort study utilized 2019 data from the National Inpatient Sample (NIS), encompassing over seven million hospital stays and covering more than 97% of the U.S. population. We compared two groups: uninsured patients aged 63-64 and Medicare-insured patients aged 65-66. The analysis focused on nine common elective surgical procedures, employing a regression discontinuity design to minimize confounding. Patients were exactly matched on several variables, including reason for hospitalization, sex, race, and hospitalization type (elective vs. emergent).

**Results:** The matched cohort, after exact matching, included 70,916 admissions (47,262 from Medicare patients aged 65-66 and 23,654 from uninsured patients aged 63-64). The study found significantly higher utilization rates of elective procedures, including cataract surgery, glaucoma surgery, joint replacement, and spinal fusion, among the Medicare-eligible group compared to the younger, uninsured group. Rates of elective procedures approximately doubled post-eligibility for Medicare. Additionally, being below the Medicare eligibility age was associated with decreased odds of undergoing these procedures and increased odds of hospital transfers.

**Discussion:** Lowering the Medicare eligibility age could lead to increased access to elective medical procedures for late middle-aged individuals, potentially reducing delays in treatment and associated complications.

## Introduction

In the United States, Medicare is the public insurer that covers all adults ages 65 and over, with some additional eligibility carve outs for people with disabilities under 65. Before this age, individuals rely on their employers or the private insurance market to obtain health coverage, yet affordable coverage near this age (51 and older) can be difficult to access, especially for those who have transitioned out of employer-sponsored health insurance or have existing health conditions. 13-25% of late middle-age individuals have been uninsured or have experienced a gap in insurance coverage in the years before obtaining Medicare.^1^ One study using data from Florida and North Carolina found large discontinuities in health care utilization for elective procedures concentrated around the Medicare eligibility threshold age of 65.^2^ As a result, policymakers have considered lowering the eligibility age for Medicare in order to expand health insurance enrollment.^3^

Lowering the eligibility age for Medicare could substantially increase the number of Americans with insurance. For example, lowering the age to 62 could increase the number of beneficiaries by over 3.9 million.^4^ However, the health impacts of such a policy have not been fully explored. Some studies found that health insurance is associated with improved health outcomes such as reduced mortality or improved treatment of chronic conditions^5^, but other studies have found no association with improved health status.^6^ The Oregon Experiment found increases in utilization of healthcare services, but no significant improvements in measured physical health outcomes.^7^ While these studies provide some insight into the potential effects of lowering the eligibility age for Medicare, they were performed at the state or interstate level. Given the heterogeneity of procedure rates and conditions across the country, a national scale is warranted to broadly investigate the association of Medicare enrollment and healthcare utilization.

Prior work has shown a significant increase in the number of hip and knee replacements performed at ages 65 and 66, as opposed to ages 63 and 64, indicating a significant increase in elective procedures performed after the introduction of Medicare.^8^ Delays in elective surgery have been associated with increased complications including infections and advanced stages of disease as well as increased costs to the system and decreased quality of life.^9^ Additionally, some studies have shown that when compared to privately or publicly insured individuals, uninsured individuals were more likely to be transferred to another hospital after being admitted to the ED.^10^

Therefore, we aimed to quantify the differences in elective procedure utilization that may result from lowering the age of Medicare eligibility to below 65. We compared 63-64 year old patients without insurance and 65-66 year old patients with Medicare using the National Inpatient Sample, investigating several outcomes including rates of elective orthopedic and ophthalmologic surgeries, the most common elective surgeries in older adults^11^, and rates of inpatient transfers out. We used a regression discontinuity approach to compare people slightly above and below the age of Medicare eligibility to reduce potential confounding from unobserved variables. We hypothesized that 65-66 year old patients on Medicare would experience increased rates of elective procedures compared to 63-64 year old patients without insurance.

## Methods

### Data source and study population

Our study used data from the National Inpatient Sample (NIS), a large all-payer inpatient care database containing data on more than seven million hospital stays in the United States (www.hcup-us.ahrq.gov/nisoverview.jsp), covering more than 97 percent of the U.S. population. The NIS is sampled from the State Inpatient Databases, including all inpatient data that are currently contributed to the Healthcare Cost and Utilization Project. This database is de-identified as state and hospital identifiers are removed. Institutional Review Board approval is not required under the Health Insurance Portability and Accountability when this dataset is used for analysis (https://www.hcup-us.ahrq.gov/DUA/dua_508/DUA508version.jsp).

We considered all admissions in 2019 for uninsured patients ages 63-64 and all patients insured by Medicare ages 65-66. We restricted our dataset to 2019 because it was the only year to include the service line variable describing reasons for hospitalization, which was integral for our exact matching approach described below. The final analytic cohort included all admissions for uninsured patients ages 63-64, and admissions for Medicare patients ages 65-66 after performing exact matching (described below; removed n = 406,701), yielding a final cohort size of 70,916 admissions.

### Outcomes

To examine the association between Medicare coverage and utilization of elective procedures, we considered nine common surgical procedures: spinal fusion, any joint replacement, cataract surgery, glaucoma surgery, bariatric surgery, liver transplant, kidney transplant, appendectomy, and cholecystectomy. A complete list of ICD and DRG codes used to identify these procedures is included in the Supplementary Information. Separate binary outcomes were defined for each of these procedures indicating whether or not the procedure occurred during a given admission. With the exception of appendectomy and cholecystectomy, the other procedures are among the most common elective procedures represented in the NIS. We hypothesized that patients in the uninsured, 63-64 age group would be less likely to receive these procedures compared to the Medicare-insured, 65-66 age group. Appendectomy and cholecystectomy were considered negative controls — since these are typically emergent procedures, we hypothesized that utilization of these two procedures would be similar between the two comparison groups.

### Exposure

We compared two groups: admissions for uninsured, Medicare-ineligible patients ages 63-64 (uninsured, 63-64); and admissions for Medicare-insured patients ages 65-66 (Medicare, 65-66). We restricted the former group to uninsured patients to understand the potential impact of expanding Medicare eligibility below the age of 65, as uninsured patients may be more likely to wait until age 65 to utilize healthcare compared to insured patients below the age of 65. We restricted the age range to two years above and below the eligibility cutoff of 65 to maximize the validity of a regression discontinuity design, where age 65 is treated as the random cutoff and patients immediately on either side of the cutoff are most likely to be similar on key confounding variables.

### Covariates

We selected the following patient variables for covariate matching: reason for hospitalization (to account for differences in care received), sex, race, and elective vs. emergent hospitalization.

### Statistical Analyses

Admissions in the treatment group were matched to admissions in the control group as follows. We performed exact matching on the following variables to compare these two groups accounting for possible confounders: For every 63-64 year-old uninsured patient, we selected two 65-66 Medicare patients. If more than two patients were eligible matches, two were selected at random. Covariate balance was confirmed through calculation of descriptive statistics. We were able to match 70,916 admissions: 47,262 from Medicare patients aged 65-66) and 23,654 from uninsured patients aged 63-64.

We calculated summary statistics comparing the matched uninsured and Medicare populations. We also plotted the various outcomes as a function of age to highlight potential discontinuities that occur at the age of Medicare eligibility (65). Lastly, we fit logistic or linear regression models to quantify the effect of Medicare eligibility on the outcomes of interest. Due to the large sample size of our cohort, hypothesis tests with a p-value of < 0.05 were not necessarily considered statistically significant since our sample was highly powered. Rather, statistical significance was considered as a continuum, and results were interpreted giving more weight to tests yielding the smallest p-values.

To examine the impact of lowering the medicare enrollment age to below 65 years old on elective procedures and hospital transfers, we used a regression discontinuity design on a retrospective cohort using inpatient data from the National Inpatient Sample (NIS) from the calendar year 2019. After filtering for patients aged 63-64 and uninsured, or patients aged 65-66 and on Medicare, and excluding stays with missing values on key variables, our analytic sample included 477,617 unique in-patient stays.

## Results

### Characteristics of the matched cohort

Before matching, the unmatched cohorts differed on several demographic characteristics. For example, in the unmatched cohorts, 45% of young, self-pay patients were female, compared to 41% of old, Medicare matched patients. Similarly, 21% of young, self-pay patients were Black, compared to 16% of old, Medicare matched patients. After 2:1 exact matching, the two groups were well-balanced on demographic characteristics, including gender, race, urban/rural location, and income quartile: 43% of patients in both groups were female, 18% were Black, 17% were from rural areas, and 40% were in the lowest income quartile. These findings suggest that matching was effective in controlling for potential confounders.

### Utilization of elective procedures and transfers out

There were significantly higher utilization rates in the Medicare-age group across all studied outcomes: transfer out, appendectomies, cataract surgery, glaucoma surgery, joint replacement surgery, and spinal fusion (Table 2). Figure 1 displays the trends graphically by showing a visual difference in mean rate in utilization or incidence between pre-65 and post-65 Medicare-eligible patients. Across most outcomes, the rates double. Being below the Medicare-eligibility age of 65 is associated with an 0.344 (95% CI: 0.307, 0.384) decreased odds of transfer outs, a 0.278 (95% CI: 0.115, 0.573) decreased odds of cataract surgery, a 0.501 (95% CI: 0.340, 0.718) decreased odds of glaucoma surgery, a 0.545 (95% CI: 0.468, 0.632) decreased odds of joint procedures, and a 0.654 (95% CI: 0.537, 0.792) decreased odds of spinal fusion (Table 3).

**Table 1.** Comparison of selected matching covariates between 63-64 uninsured (“Young, Self-pay’’), 65-66 Medicare matched (“Old medicare, matched”), and 65-66 Medicare unmatched (“Old medicare, unmatched”).

| **Characteristic** | **Young, Self-pay, N =**<br>**23,654<sup>†</sup>** | **Old medicare, matched, N**<br>**= 47,262<sup>†</sup>** | **Old medicare, unmatched, N =**<br>**453,963<sup>†</sup>** |
| --- | --- | --- | --- |
| % Female | 10,182 (43%) | 20,358 (43%) | 228,318 (50%) |
| Race |  |  |  |
| API | 507 (2.1%) | 1,013 (2.1%) | 8,670 (1.9%) |
| Black | 4,302 (18%) | 8,603 (18%) | 65,117 (14%) |
| Hispanic | 3,593 (15%) | 7,162 (15%) | 32,922 (7.3%) |
| Native American | 124 (0.5%) | 240 (0.5%) | 2,605 (0.6%) |
| Other | 862 (3.6%) | 1,715 (3.6%) | 9,789 (2.2%) |
| White | 14,266 (60%) | 28,529 (60%) | 334,860 (74%) |
| Urban/Rural |  |  |  |
| Rural | 3,938 (17%) | 7,848 (17%) | 83,530 (18%) |
| Urban | 19,716 (83%) | 39,414 (83%) | 370,433 (82%) |
| Income quartile |  |  |  |
| 1 | 9,562 (40%) | 19,116 (40%) | 142,780 (31%) |
| 2 | 6,269 (27%) | 12,523 (26%) | 118,683 (26%) |
| 3 | 4,860 (21%) | 9,699 (21%) | 110,914 (24%) |
| 4 | 2,963 (13%) | 5,924 (13%) | 81,586 (18%) |

**Table 2.** Comparison of outcomes between 63-64 uninsured (“Young, Self-pay”) and 65-66 Medicare matched (“Old medicare, matched”).

| **Characteristic** | **Old medicare, matched, N =**<br>**27,874<sup>1</sup>** | **Young, Self-pay, N =**<br>**13,587<sup>1</sup>** | **p-value<sup>2</sup>** |
| --- | --- | --- | --- |
| Transfer out | 6,611 (24%) | 1,292 (9.5%) | <0.001 |
| Appendectomy | 93 (0.3%) | 108 (0.8%) | <0.001 |
| Cataract | 210 (0.8%) | 27 (0.2%) | <0.001 |
| Glaucoma | 512 (1.8%) | 87 (0.6%) | <0.001 |
| Joint<br>replacements | 3,702 (13%) | 655 (4.8%) | <0.001 |
| Spinal fusion | 2,345 (8.4%) | 408 (3.0%) | <0.001 |

<sup>1</sup> n (%)
<sup>2</sup> Pearson's Chi-squared test

**Table 3.** Odds ratios with 95% confidence intervals from regression discontinuity modeling for each of the outcomes. The outcome is modeled with the reference group as the “Old, Medicare matched”.

| Outcome Variable | OR (95% CI) | P-value |
| --- | --- | --- |
| Transfer Out | 0.344 (0.307, 0.384) | <0.001 |
| Appendectomy | 1.15 (0.730, 1.77) | 0.54 |
| Cataract Surgery | 0.278 (0.115, 0.573) | 0.0015 |
| Glaucoma Treatment | 0.501 (0.340, 0.718) | <0.001 |
| Joint Procedure | 0.545 (0.468, 0.632) | <0.001 |
| Spinal Fusion | 0.654 (0.537, 0.792) | <0.001 |

**Figure 1.**
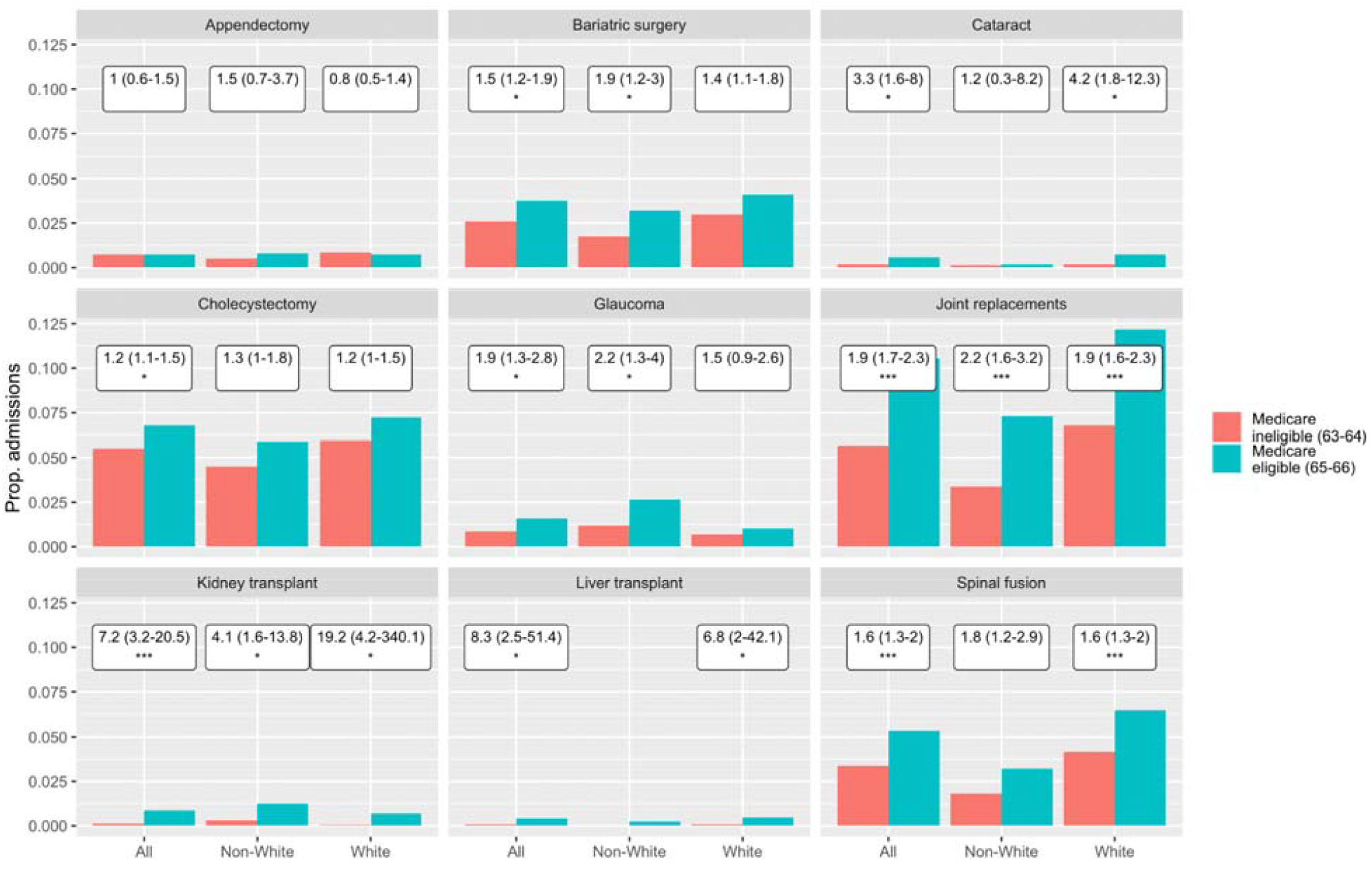
For each outcome, proportion of inpatient admissions resulting in the outcome of interest for Medicare ineligible (red) and Medicare eligible (blue). Odds ratios and 95% CI are shown above each bar. * = p<0.01, ** = p<0.001, *** = p<0.0001.

## Discussion

In this retrospective cohort study of 70,916 inpatient hospitalization records from the NIS in 2019, we found that patients from 65 to 69 years old with Medicare utilized significantly more health system services compared to patients from 60 to 64 years old without insurance for the same medical condition. Patients with Medicare had higher rates of elective orthopedic and ophthalmologic procedures and higher rates of transfers out of inpatient services.

A scoping review shows that Medicare enrollment was found to increase self-reported health and healthcare utilization, as well as reduce racial and socioeconomic disparities. Our results show that elective surgery rates were significantly higher in the 65-66 year old patient group. This corroborates previous findings of increased healthcare usage after age 65 among previously uninsured adults.^12^It’s possible that the costs of expanding coverage for uninsured near-elderly adults with these conditions may be partially offset by subsequent reductions in health care use and expenditures after they reach age 65.

Our study indicates a difference in elective surgery utilization rates for individuals upon enrollment in Medicare. This has important implications for insurance coverage for those who are approaching the Medicare enrollment age of 65. Our findings suggest policy makers should consider health policy levers such as subsidies on premiums for the near-elderly, a new purchasing arrangement to lower premiums, a State Medicare buy-in, or lowering the age for Medicare eligibility to expand access to Medicare.

Our analysis has some limitations, including the lack of longitudinal analysis for the use of health services, physician cost, and health outcomes due to the nature of the NIS dataset. The observational design did not allow us to make definitive conclusions about the causal effects of acquiring health insurance on subsequent reductions in health care use or expenditures. With regards to mortality, there may be confounding variables with the uninsured dying at higher rates outside the hospital and therefore not being accurately captured. Some variables may have been better served by data disaggregation, such as ‘Asian race’ rather than a breakdown by specific racial category. Additionally, we did not consider subgroups based on diagnosis— differences in cost and mortality may be observed when focusing on a particular diagnosis group. We could have also examined the racial distribution of the patients who are uninsured and on Medicare, which would add to the urgency in which we should implement policy reforms to expand Medicare to increase equity.

For instance, studies have had limitations assessing in-hospital mortality differences between non-Medicare insured and recently Medicare insured patients due infrequent deaths reported or short evaluation periods masking improvements in health. Additionally, There is sparse literature examining the nationwide impact of Medicare enrollment on healthcare utilization, particularly after the implementation of the Affordable Care Act. This is particularly relevant as Medicare reimbursements are lower than private insurance, so more medicare beneficiaries may not equal more healthcare utilization or better outcomes if providers do not accept these patients.

One especially challenging aspect to studying the potential effect of Medicare expansion is unobserved differences between persons with and without insurance, which could confound simple comparisons between people with and without insurance.

## Data Availability

Data used for this analysis is available from HCUP at www.hcup-us.ahrq.gov/nisoverview.jsp.

https://www.hcup-us.ahrq.gov/nisoverview.jsp.

## Supplementary Information

**Supplementary Table 1:**
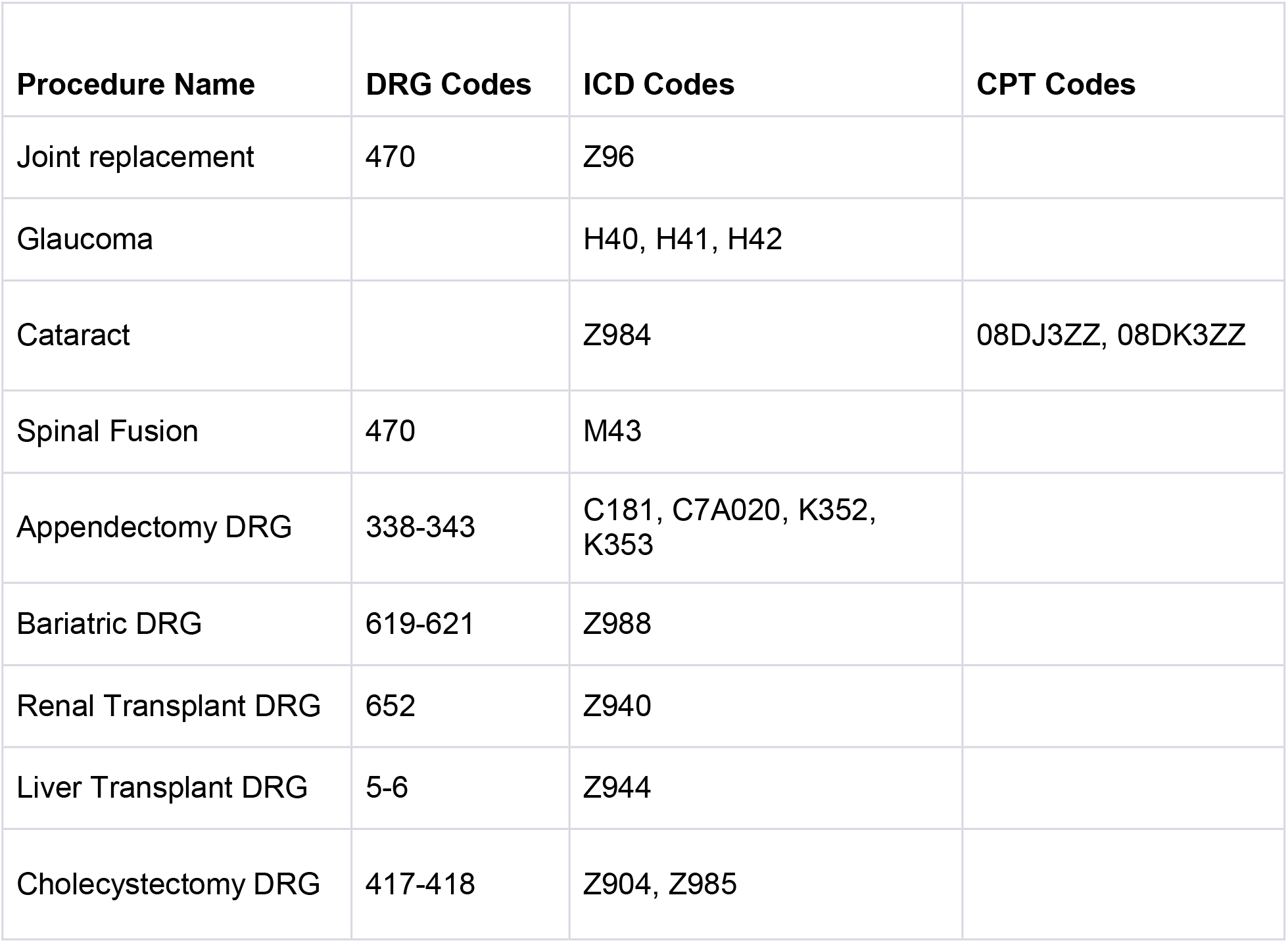
DRG, ICD, and CPT codes used to define the outcomes of interest.

## References

1. Baker DW, Sudano JJ. Health insurance coverage during the years preceding medicare eligibility. Arch Intern Med. 2005 Apr 11;165(7):770–6.

2. David G, Saynisch P, Acevedo-Perez V, Neuman MD. Affording to wait: Medicare initiation and the use of health care. Health Econ. 2012 Aug;21(8):1030–6.

3. Rep. Jayapal P [D W 7. Text - H.R.5165 - 117th Congress (2021-2022): Improving Medicare Coverage Act [Internet]. 2021 [cited 2023 Sep 18]. Available from: https://www.congress.gov/bill/117th-congress/house-bill/5165/text

4. Holt C, Parente S. Lowering the Medicare Age to 60: Cost and Coverage Outcomes [Internet]. AAF. [cited 2023 Sep 18]. Available from: https://www.americanactionforum.org/research/lowering-the-medicare-age-to-60-cost-and-coverage-outcomes/

5. Sommers BD, Gawande AA, Baicker K. Health Insurance Coverage and Health — What the Recent Evidence Tells Us. N Engl J Med. 2017 Aug 10;377(6):586–93.

6. Polsky D, Doshi JA, Escarce J, Manning W, Paddock SM, Cen L, et al. The Health Effects of Medicare for the Near-Elderly Uninsured. Health Serv Res. 2009;44(3):926–45.

7. Baicker K, Taubman SL, Allen HL, Bernstein M, Gruber JH, Newhouse JP, et al. The Oregon Experiment — Effects of Medicaid on Clinical Outcomes. N Engl J Med. 2013 May 2;368(18):1713–22.

8. Rankin KA, Gibson D, Schwarzkopf R, O’Connor MI, Wiznia DH. Operative Techniques to Reduce Hip and Knee Arthroplasty Complications in Morbidly Obese Patients. Arthroplasty Today. 2022 Aug 29;17:120–5.

9. Fu SJ, George EL, Maggio PM, Hawn M, Nazerali R. The Consequences of Delaying Elective Surgery: Surgical Perspective. Ann Surg. 2020 Aug;272(2):e79–80.

10. Venkatesh AK, Chou SC, Li SX, Choi J, Ross JS, D’Onofrio G, et al. Association Between Insurance Status and Access to Hospital Care in Emergency Department Disposition. JAMA Intern Med. 2019 May 1;179(5):686–93.

11. Deiner S, Westlake B, Dutton RP. Patterns of Surgical Care and Complications in the Elderly. J Am Geriatr Soc. 2014 May;62(5):829–35.

12. McWilliams JM, Meara E, Zaslavsky AM, Ayanian JZ. Use of health services by previously uninsured Medicare beneficiaries. N Engl J Med. 2007 Jul 12;357(2):143–53.

